# Monitoring the rise of the SARS-CoV-2 lineage B.1.1.7 in Tenerife (Spain) since mid-December 2020

**DOI:** 10.1101/2021.03.14.21253535

**Authors:** Julia Alcoba-Florez, Jose M. Lorenzo-Salazar, Helena Gil-Campesino, Antonio Íñigo-Campos, Diego García-Martínez de Artola, Victor García-Olivares, Oscar Díez-Gil, Agustín Valenzuela-Fernández, Laura Ciuffeda, Rafaela González-Montelongo, Carlos Flores

## Abstract

Starting in December 2020, a sharp increase of COVID-19 cases occurred in Tenerife compared to the rest of the Canary Islands (Spain). Because of the direct touristic connections between Tenerife and the UK, and the rapid transmission and dominance of the SARS-CoV-2 B.1.1.7 variant of concern (VOC-202012/01) by the end of November 2020 in South England, here we measured the proportion of B.1.1.7 cases occurring between the 18^th^ of December 2020 and the 25^th^ of February 2021. Out of the 2,091 COVID-19 positive nasopharyngeal swab samples assessed, 226 showed a spike gene target failure (SGTF). Subsequent viral genome sequencing further confirmed that 93.2% of them corresponded to the B.1.1.7 lineage. Furthermore, a rapid increase in the proportion of SGTF variants was detected in up to 10.7% of positive cases during the Christmas season despite stricter measures for containing the transmission were imposed in Tenerife in the period. These results support the local transmission of SARS-CoV-2 B.1.1.7 lineage in Tenerife since late December 2020 although it is not yet dominant.

## Introduction

Tenerife (Canary Islands, Spain) is one of the top destinations for UK travelers all year long and was an air corridor until late 2020 (Department for Transport and Foreign, Commonwealth & Development Office 2020) as the coronavirus disease 2019 (COVID-19) had <50 cases of accumulated incidence per 100,000 inhabitants in the last 14 days (AI14) until late September, retaining one of the lowest case levels among all Spanish communities since the decrease of the first wave during April 2020 (Ministerio de Sanidad 2020). Starting in December 2020, Tenerife faced an unexplained rapid increase of COVID-19 cases compared to the rest of the Archipelago as AI14 jumped from <130 cases between August and late November 2020 to 248 cases by the 23^rd^ of December 2020 (https://www.gobiernodecanarias.org/principal/coronavirus/acceso_datos.html). As a consequence, the authorities imposed border restrictions and recommended limiting the mobility between municipalities from the 19^th^ of December 2020 until the 2^nd^ of January 2021.

The emergence of novel Severe Acute Respiratory Syndrome Coronavirus 2 (SARS-CoV-2) variants is generating widespread concern, as they harbor a constellation of mutations that might lead to increased transmissibility (Kirby 2021) and/or immune evasion from previous infection or vaccination (Wu et al. 2021). In particular, the lineage B.1.1.7 (variant of concern [VOC]-202012/01 according to Public Health England; also known as clade 20I/501Y.V1), firstly identified in the UK in December 2020 (O’Toole et al. 2021), has rapidly spread worldwide, with 82 countries reporting its detection by the 15^th^ of February (https://cov-lineages.org/global_report_B.1.1.7.html). By mid-January 2021, this VOC had an accumulated prevalence over 76% among all UK sequences despite restrictions in many of the affected areas (O’Toole et al. 2021). Among the mutations characterizing this lineage, nine are found in the spike (S) protein gene compared to the reference genome (SARS-CoV-2 isolate Wuhan-Hu-1; GenBank accession MN908947). Within these, the deletion of the 21765-21770 genome region that predicts the loss of the amino acids 69 and 70 (Δ69/70) in the S protein has been associated with a “Spike gene target failure” (SGTF) for two commercial RT-qPCR kits for COVID-19 diagnosis (Public Health England 2020; FDA COVID-19 Update, 2021). While this characteristic is an imperfect proxy, it has helped tracking the rapid emergence of the B.1.1.7 lineage around the world (Washington et al. 2020). Here we aimed to monitor the SARS-CoV-2 B.1.1.7 lineage cases in Tenerife beginning in mid-December 2020 by leveraging the SGTF during the RT-qPCR followed by viral genome sequencing of the prioritized samples.

## Materials and Methods

The study was conducted at the University Hospital Nuestra Señora de Candelaria (Santa Cruz de Tenerife, Spain). We assessed nasopharyngeal swabs from COVID-19/SARS-CoV-2 patients corresponding to independent outbreaks declared from 18^th^ of December 2020 to 25^th^ of February 2021. Routine COVID-19 testing in the center was conducted using diverse commercial RT-qPCR alternatives, as described elsewhere (Alcoba-Florez et al. 2020).

We used the TaqPath COVID-19 Combo Kit (Thermo Fisher Scientific) to monitor the drop out in the S viral gene (one of the three gene targets of the kit) in the COVID-19 positive patients. This SGTF has been suggested to occur by the presence of the Δ69/70 deletion (FDA COVID-19 Update, 2021) that is known to have arisen in multiple lineages (Davies et al. 2021). We used this assay to assess all RNA extracts (Zymo Quick DNA/RNA kit, Zymo) from COVID-19 positives in the period as an imperfect proxy of the incidence of the B.1.1.7 lineage in the population, resulting in a reassessment of a total of 2,091 positive samples in the period. Thermocycling was conducted on a 7500 Fast Real-Time PCR System (Thermo Fisher Scientific) following manufacturers recommendations.

Samples were selected for sequencing if they showed a cycle threshold (Ct)<30 for any of the targets included in the COVID-19 diagnosis kits and were categorized as SGTF. The libraries were obtained using the COVIDSeq Test (Illumina, Inc.), with a proven high reproducibility and a sensitivity comparable to that of RT-qPCR assays (Bhoyar et al. 2021), covering the whole SARS-CoV-2 genome with 98 amplicons and including internal controls consisting of 11 human mRNA targets. The procedure followed the manufacturers recommendations and quality control steps. Sequencing was conducted on a NextSeq 550 (Illumina) instrument on High Output mode with 36 bp single end reads at the Instituto Tecnológico y de Energías Renovables (Santa Cruz de Tenerife, Spain).

Per sample consensus sequences and variants against the reference (NC_045512.2) were obtained based on the DRAGEN COVIDSeq Test v1.2.2 pipeline (Illumina, Inc.). Positive and negative amplification controls were included in each run. To be able to recover the FASTA sequences that were not automatically provided by DRAGEN COVIDSeq Test, the fastq files were reprocessed with DRAGEN Lineage v3.5.0 with the following specifications: coverage threshold of 20, virus detection threshold of 5, virus negative threshold of 5, and a human control threshold of 3. Phylogenetic Assignment of named Global Outbreak LINeages (PANGOLIN) software suite v2.1.7 (https://github.com/cov-lineages/pangolin) was then used for lineage and clade classification following the nomenclature of Rambaut et al. (2020). Nextclade v.0.12.0 (Hadfield et al., 2018) was used for variant calling and functional predictions.

The AI14 data was retrieved from the COVID-19 data hub from Gobierno de Canarias (https://www.gobiernodecanarias.org/principal/coronavirus/acceso_datos.html; accessed March 6^th^, 2021).

## Results

We closely followed SGTF of the SARS-CoV-2 samples in Tenerife in the period of 2 months (18^th^ of December 2020 to 25^th^ of February 2021). A total of 2,091 COVID-19 positive samples were assessed in the period, resulting in 226 SGTF samples identified and reaching a maximum of 10.7% at the end of the period under study. Remarkably, we detected a rapid increase in the prevalence of SGTF over this period, rising sharply during the Christmas season and steadily after December 28^th^, 2020 (**Figure 1**).

**Fig. 1.**
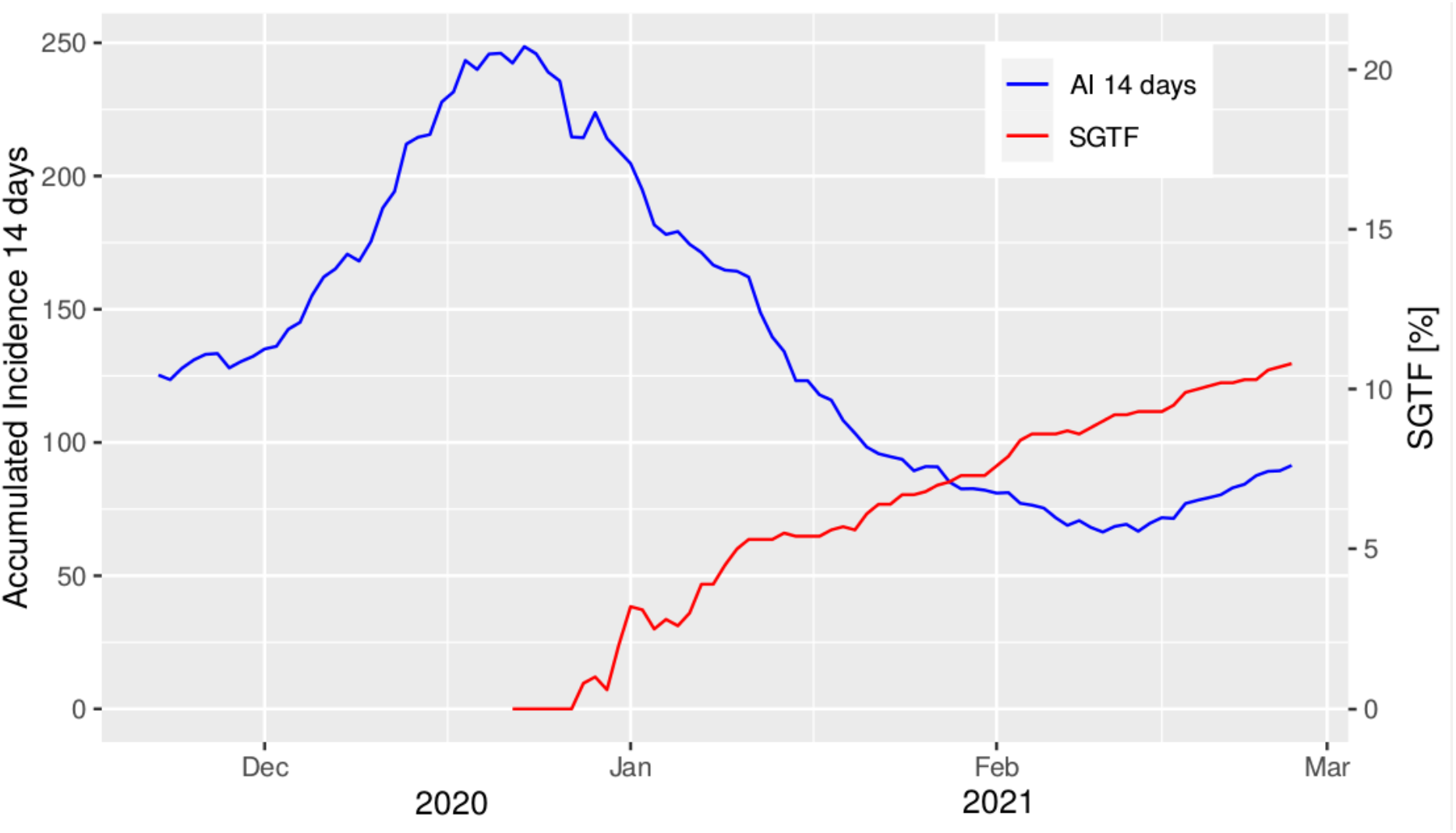
Accumulated incidence per 100,000 inhabitants in the last 14 days (AI 14 days) and spike gene target failure (SGTF) cases per day in the study period.

We were able to sequence 147 (mean depth of coverage ± SD: 48X ± 9) out of the 226 SGTF samples in the period. The sequenced genomes of SARS-CoV-2 with SGTF were assigned to five different lineages where B.1.1.7 (VOC-202012/01) was the predominant (93.2%). Eleven of these had lower sequence quality (according to the quality control assessment carried out by Nextclade based on the aggregation of four metrics: missing data, mixed sites, private mutations, mutation clusters) and were not included in downstream analysis. Among those classified as B.1.1.7 lineage, the sequences carried between nine and 17 of the lineage-defining mutations (Davies et al. 2021), with 93.4% of the sequences including ≥15 (**Figure 2**). Other lineages that were linked to the SGTF in this study were classified as B.1.258 (5), B.1.177 (3), B.1.221 (1), and B.1.1.222 (1).

**Fig. 2.**
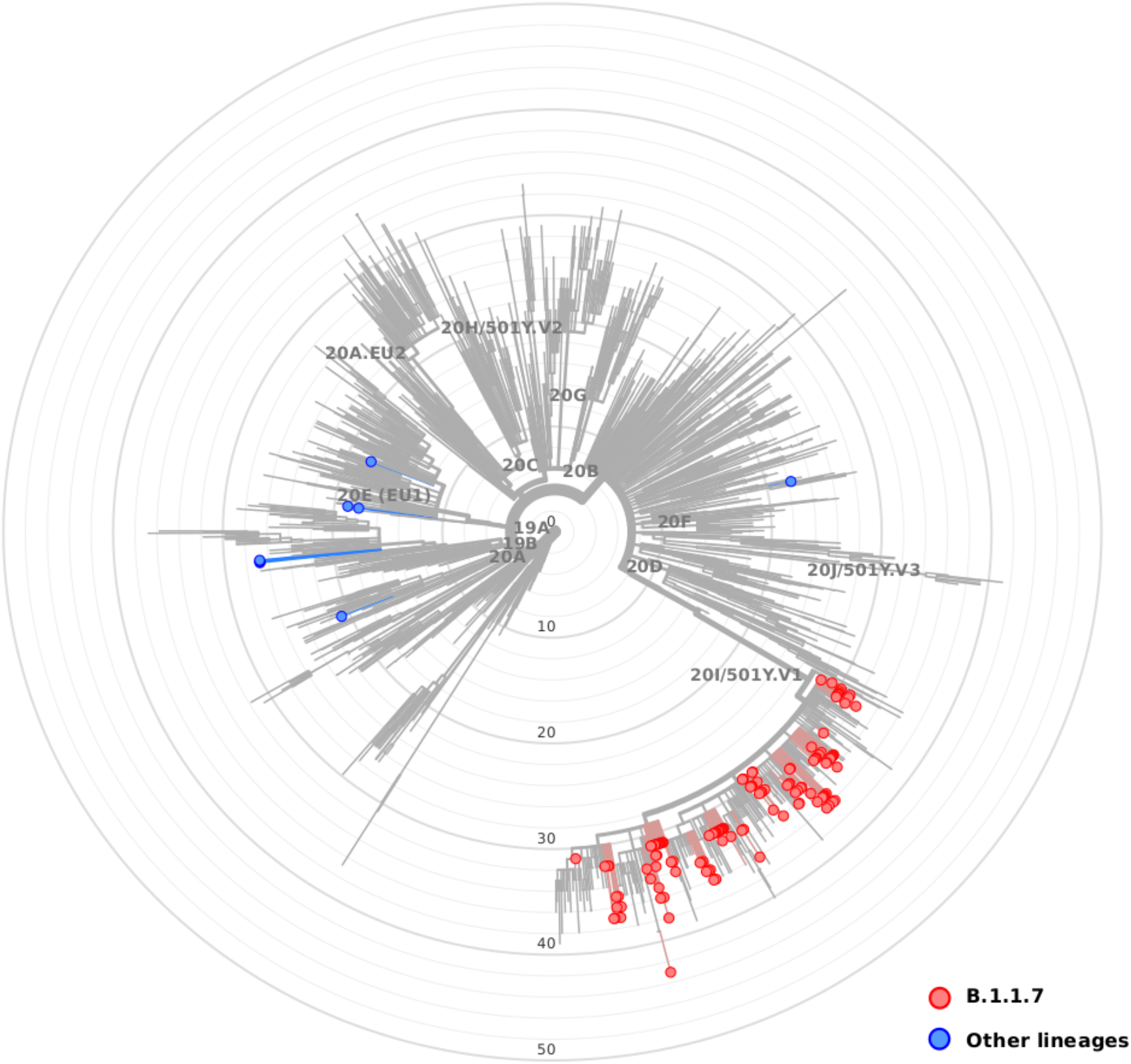
Maximum likelihood phylogenetic tree of SARS-CoV-2 sequences characterized by the spike gene target failure (SGTF) in the period (B.1.1.7 lineages in red; other lineages in blue). The tree also includes relationships with other 1,902 genome sequences representative of major clades, labelled based on Nextstrain definitions, from Auspice v2.23.0 (as of 2021-03-07) and sampled between December 2019 and March 2021 for reference (light grey).

## Discussion

Based on RT-qPCR SGTF and viral genome sequencing, this study supports the local transmission of SARS-CoV-2 B.1.1.7 lineage in Tenerife since late December 2020. Despite vaccination campaign started in Spain on the 27^th^ of December 2020 and given that it has not reached a large fraction of the population yet, the possibility that the B.1.1.7 lineage is also responsible for increased transmission in the rest of the Archipelago is high. Thus, a denser analysis is likely to detect further spread of the B.1.1.7 lineage across the Canary Islands, especially if sampling is conducted after the 12^th^ of February 2021 when the mobility increased again in all islands except for the island of Lanzarote.

Our observations for the island of Tenerife support that B.1.1.7 is not yet dominant as of 25^th^ of February 2021. Although neglecting a protective effect from vaccination, the models for some major cities in Europe indicate that the B.1.1.7 lineage will be the dominant (>50% of positives) by mid-March 2021 even if accounting for non-pharmaceutical interventions (Gozzi et al. 2021). Therefore, the SARS-CoV-2 B.1.1.7 lineage could be dominant also in Tenerife during the first half of 2021.

We anticipate that the data reported here should be taken as an underestimate as there will be genomes of the B.1.1.7 lineage that escape detection by RT-qPCR SGTF. We also assume that there is potential that some of the samples may derive from non-resident travelers that could have introduced an upward bias in the estimated AI14.

## Data Availability

Data will be publicly available through GISAID.

## Author contributions

JAF and CF designed the study. JAF, JMLS, HGC, DGM, RGM, AIC, VGO, and ODG participated in data acquisition. JMLS and CF performed the analyses and data interpretation. JMLS, AVF, LC, RGM, and CF wrote the draft of the manuscript. All authors contributed to the critical revision and final approval of the manuscript.

## Acknowledgements

We deeply acknowledge the University Hospital Nuestra Señora de Candelaria and the Instituto Tecnológico y de Energías Renovables board of directors for their strong support and assistance in accessing diverse resources used in the study.

## Ethical Approval

The University Hospital Nuestra Señora de Candelaria (Santa Cruz de Tenerife, Spain) review board approved the study (ethics approval number: CHUNSC_2020_24).

